# Hospital practice in COVID-19 times: perceptions of the midwifery interns in Peru

**DOI:** 10.1101/2020.06.05.20094482

**Authors:** Jessica Rojas-Silva, Valery Damacen-Oblitas, Diayan Castro-Gomez, Jennifer Rojas-Vega, John Barja-Ore, Randol Vila-Arevalo, Victor Moquillaza-Alcantara

**Author notes:** **Corresponding author: Victor Hugo Moquillaza Alcántara Address:** Condominio Los Nogales, Ed.8, Dto.503. Urb. Los Parques de El Agustino. El Agustino. Lima, Peru.; **email:**.

## Abstract

**Objective:** To evaluate the perception of midwifery interns regarding hospital practices during COVID-19.

**Material and methods:** Study of qualitative approach, of phenomenological design, where 80 midwifery interns from the different regions of Peru participated, who are also representatives of their hospital headquarters. An in-depth interview was applied where the perception of hospital practices was addressed according to: i) current problems and ii) solution proposals.

**Results:** Midwifery interns have been removed from hospital practices, mainly due to the absence of personal protective equipment and health insurance, financially affecting those who must continue to make rent and food payments; Likewise, a large part of the universities have not offered proposals for solutions to the delay in internships, raising concerns about delays in administrative procedures, even more so for students from non-licensed universities. Among the proposals, those who are close to graduating suggest being exempted from the months when there were no activities, so as not to delay future processes such as tuition and rural service; likewise, suspend payment for these months and strengthen knowledge through the discussion of clinical cases, which could be virtual.

**Conclusions:** The cessation of hospital practice responds to a lack of guarantees in the health care of the student, generating economic repercussions and a negative perception regarding university management. Finally, solutions that could be considered for the next decisions made by the institutions are reported.

## INTRODUCTION

At the end of the year 2019, in the city of Wuhan, China, the outbreak of cases with atypical pneumonia began, spreading in various countries and finally being named as “Coronavirus Disease 2019” (COVID-19) (1,2). Faced with the rapid spread of COVID-19, the World Health Organization declared a state of international health emergency (3). With which, countries from all regions of the world opted for measures that reduce the chain of contagion, among which the obligatory social isolation stands out, the same that has generated repercussions on different activities of the population (4,5).

In the educational field, this situation has led to the suspension of face-to-face academic activities and the requirement for remote education at the basic and higher levels (6). Case is presented in health science students, who part of their professional training involves in-hospital practices, being affected by the restriction of access to facilities. In this regard, guidelines have been proposed to include students in the surveillance of patients with COVID-19, for which minimum requirements must be taken into account, such as the availability of personal protective equipment and screening tests, as well as the assessment of their competences to accept responsibility, regardless of the year of study (7.8). Among other proposals is also the inclusion of the student in public health programs, with previous training in the preventive measures corresponding to COVID-19 (9).

In Peru, the state of national emergency was declared on March 16, 2020 (10), along with other policies and measures in favour of national public health. Meanwhile, the National Superintendence of University Higher Education urged that higher education institutions reprogram their academic and administrative activities in person and invoked the use of information technologies to continue university education (11). The specific situation of the training of future health professionals in the face of the recent situation is unknown, due to the lack of communication spaces between public and private universities with their student group. In addition to this, it is still uncertain how and when the process of reincorporation of midwifery interns to hospital teaching facilities will begin (12,13), especially considering the educational and social needs that each one presents in the different regions. From Peru. In this sense, this investigation was carried out with the objective of evaluating the perception of midwifery interns from the regions of Peru regarding hospital practices in the days of COVID-19.

## MATERIAL Y METHODS

### TYPE OF STUDY

Inductive qualitative approach study developed under the phenomenological paradigm, where the experience of an event is studied from the perspective of the individual, being significant in subtracting the personal perspective and its interpretation and sidestepping assumptions predisposed as true (14).

### PARTICIPANTS

Students from the last year of midwife (midwifery interns) who were appointed as representatives, also considered as “delegates”, were selected from a group of students who carry out their hospital practices. The representatives are also those who maintain communication with the coordinators or “tutors” of each hospital site, who represent the university in their institution.

The list of inmates fulfilling the role of representatives in the regions of Peru was requested from the National Association of Midwifery Students of Peru (ANEOP, in Spanish), which numbered 98 students, who were invited to participate voluntarily in the study. Finally, 80 representatives agreed to join the study, who were enrolled to participate in an in-depth personal interview remotely (online).

### VARIABLES

The student’s perception of hospital practice in a COVID-19 context was considered as the main characteristic to be evaluated. To address this perception, a guide to questions was prepared, which was made up of two sections: i) Current situation and perceived problems and ii) Proposals for solutions considered by the group they represent.

### ANALYSIS OF THE INFORMATION

The information obtained was reviewed by the researchers, selecting per student the message or messages that summarize or represent the main idea that they sought to communicate, which were transcribed in an Excel database, where each row corresponded to an midwifery representative and each column the questions asked.

During the review of the responses regarding the “current situation and perceived problems”, four recurring topics were observed, which were considered as research codes: i) Health care, ii) Economic repercussions, iii) University management (Procedures and Posture of the universities). Therefore, a thematic analysis was then carried out based on the codes finally considered.

### ETHICAL ASPECTS

The study had the review and approval of the ethics committee of the “Committee for Ethics in Specific Research for COVID-19” of the Institute for Health Technology Assessment and Research, in Lima Peru. During the investigation, it was guaranteed that all participation was voluntary and informed. Finally, the identification of the students was coded before transcription from the database.

## RESULTS

Eighty midwifery students at the national level were interviewed, who resided mainly in the Lima (22.5%) and Huancavelica (13.75%) regions. (**Table 1**) Likewise, each participant represented, on average, 12.58 ± 14.14 (Me: 8) midwifery inmates in their respective hospital centres. On the other hand, 17.50% of the interviewees reported that the hospital’s internship practices were scheduled to end in December 2020, just as 13.75% reported that this would end in November.

**Table 1.**
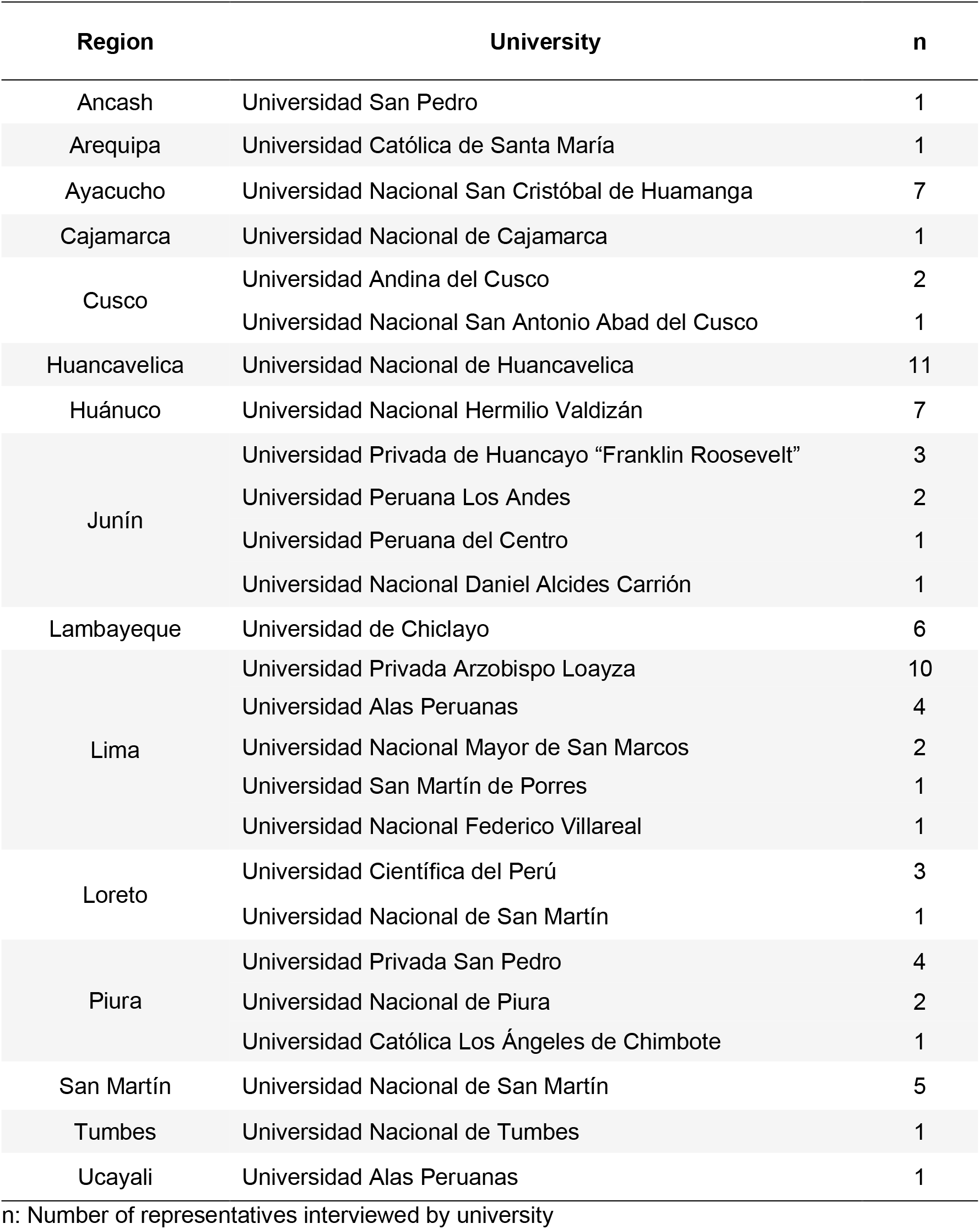
Origin of midwifery interns interviewed in Peru, 2020

| Region | University | n |
| --- | --- | --- |
| Ancash | Universidad San Pedro | 1 |
| Arequipa | Universidad Católica de Santa María | 1 |
| Ayacucho | Universidad Nacional San Cristóbal de Huamanga | 7 |
| Cajamarca | Universidad Nacional de Cajamarca | 1 |
| Cusco | Universidad Andina del Cusco | 2 |
|  | Universidad Nacional San Antonio Abad del Cusco | 1 |
| Huancavelica | Universidad Nacional de Huancavelica | 11 |
| Huánuco | Universidad Nacional Hermilio Valdizán | 7 |
| Junín | Universidad Privada de Huancayo “Franklin Roosevelt” | 3 |
|  | Universidad Peruana Los Andes | 2 |
|  | Universidad Peruana del Centro | 1 |
|  | Universidad Nacional Daniel Alcides Carrión | 1 |
| Lambayeque | Universidad de Chiclayo | 6 |
| Lima | Universidad Privada Arzobispo Loayza | 10 |
|  | Universidad Alas Peruanas | 4 |
|  | Universidad Nacional Mayor de San Marcos | 2 |
|  | Universidad San Martín de Porres | 1 |
|  | Universidad Nacional Federico Villareal | 1 |
| Loreto | Universidad Científica del Perú | 3 |
|  | Universidad Nacional de San Martín | 1 |
| Piura | Universidad Privada San Pedro | 4 |
|  | Universidad Nacional de Piura | 2 |
|  | Universidad Católica Los Ángeles de Chimbote | 1 |
| San Martín | Universidad Nacional de San Martín | 5 |
| Tumbes | Universidad Nacional de Tumbes | 1 |
| Ucayali | Universidad Alas Peruanas | 1 |

n: Number of representatives interviewed by university

Next, the problems or perceptions of the current situation of hospital practices were evaluated. **Figure 1** shows the responses obtained regarding the lack of personal protective equipment (PPE), which became constant in the establishments; likewise, they refer that the hospital authorities do not guarantee that during the return to the practices the necessary equipment will be provided, since these are only available to the professionals who work there; On the other hand, in the face of hospital exposure, they state that there is a lack of affiliation to comprehensive insurance that would respond immediately to a possible contagion.

**Figure 1.**
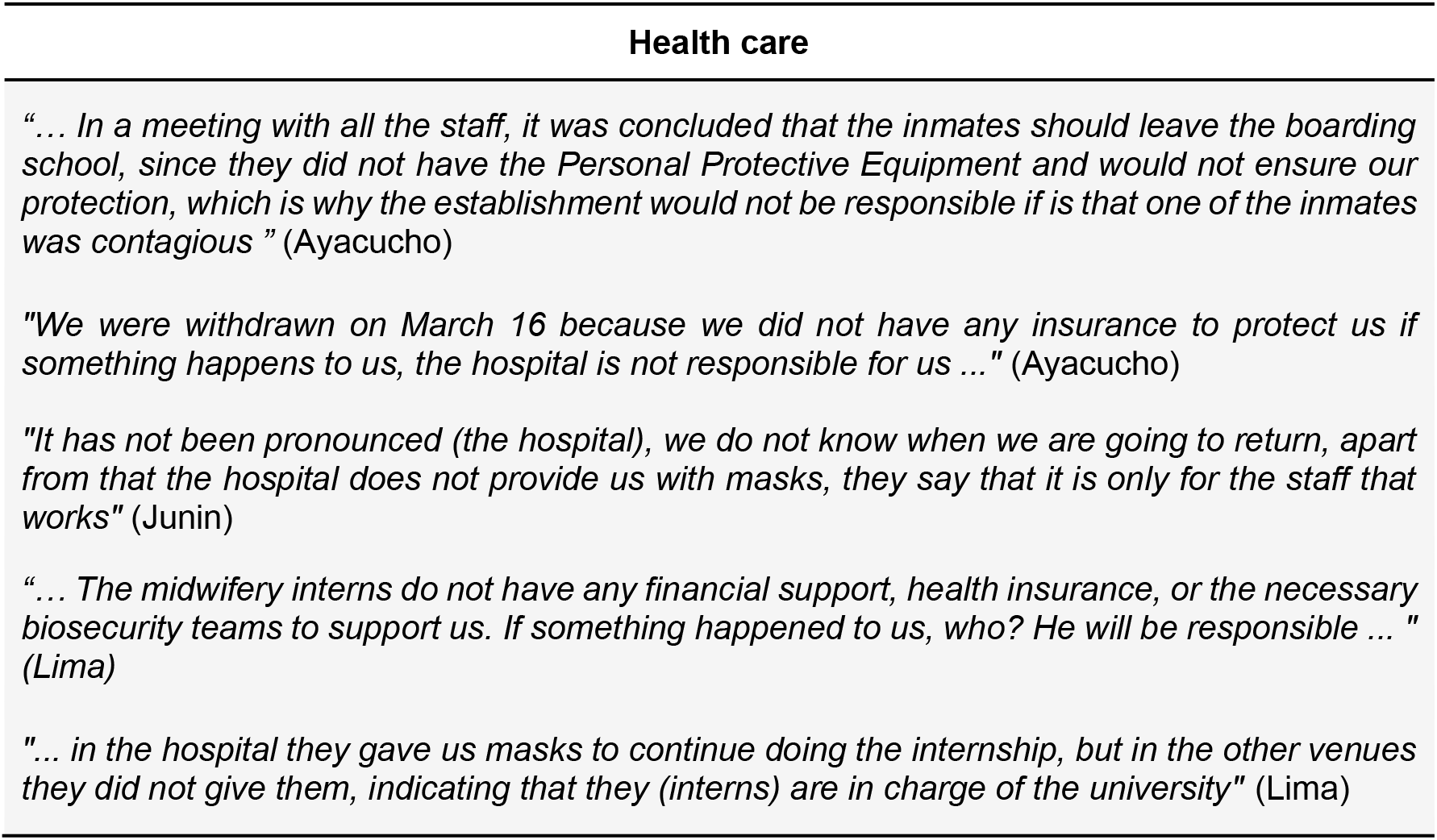
Perceived problems regarding the health care of Peruvian midwifery interns due to COVID-19

Another perceived problem was the economic repercussions, summarized in **Figure 2**. It was found that there are regions where students must travel to another province in order to complete their internship, with which they are currently obliged to continue paying for accommodation and food, despite not be doing clinical practices since the return to the boarding school is constantly postponed.

**Figure 2.**
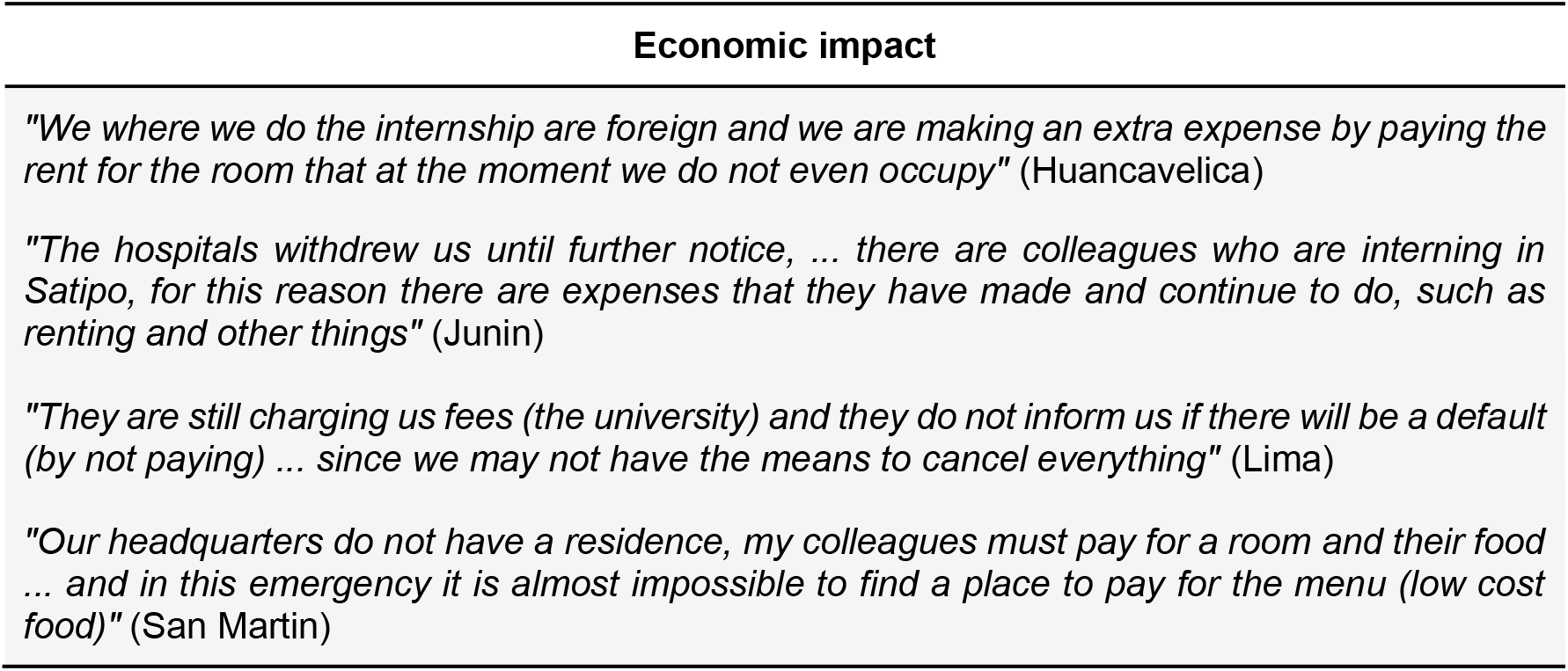
Perceived economic repercussions in Peruvian midwifery interns due to COVID-19

**Figure 3** shows the perceived problems regarding university management, among which were: i) the delay in completing certain procedures, which causes concern for students nearing graduation who require the degree to start their schooling, as well as those inmates from universities that did not obtain a university degree, and ii) the position adopted by certain institutions, who have suggested an immediate return to clinical practices, where inmates have usually preferred not to go due to lack of necessary protection and In other situations, they have been mandatory, referring that not attending their hospital rotations would be sanctioned with a suspension of their previous rotations, proposing that they begin the internship again next year.

**Figure 3.**
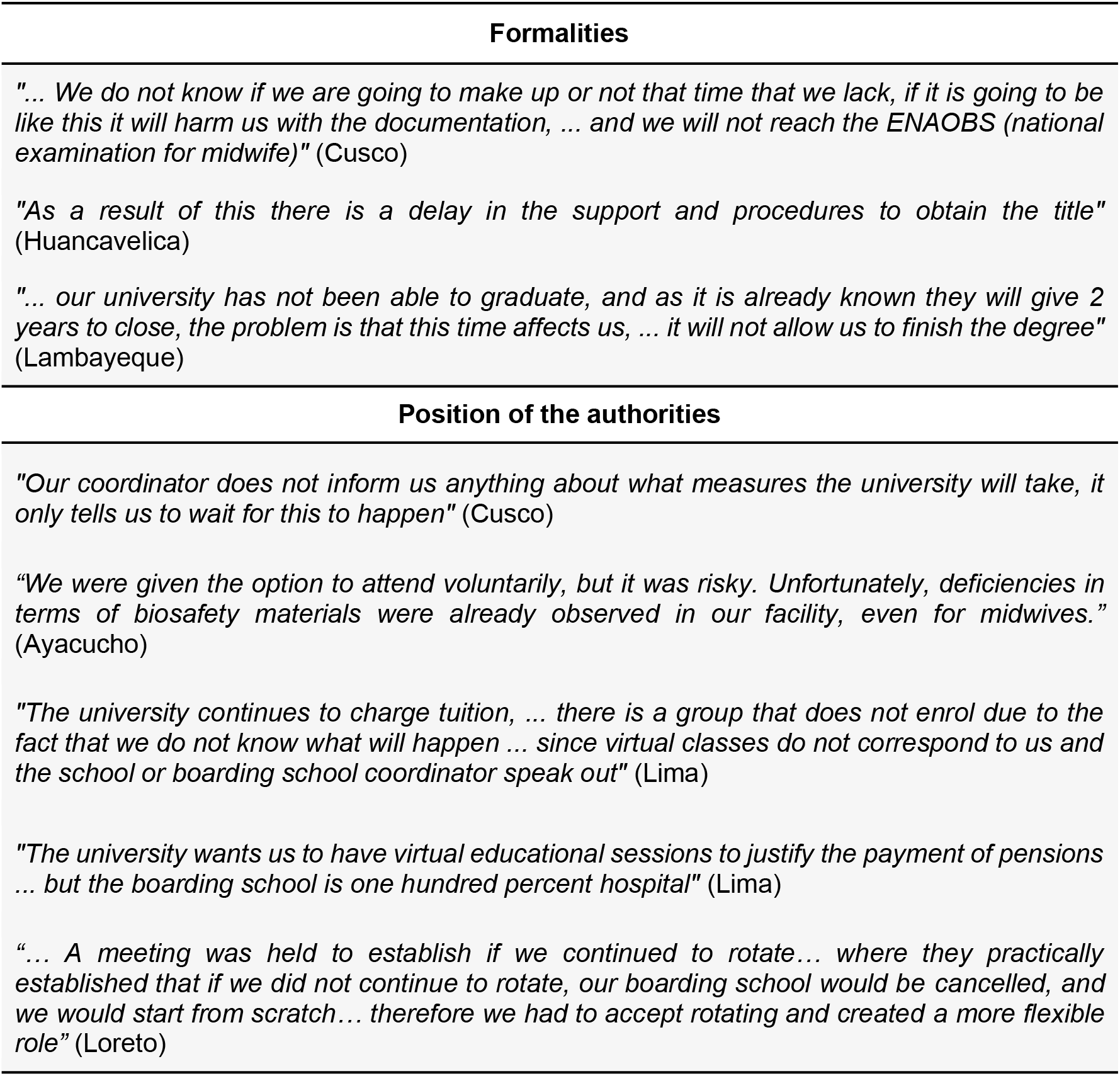
Perceived problems with university management in Peruvian midwifery interns due to COVID-19

Finally, **Figure 4** reports the comments regarding the proposed solutions by the student representatives. It was found that the students are aware that the state of emergency limits the immediate return to hospital services and that the security protocol does not include inmates, which is why they suggest the provision of PPE and the activation of comprehensive insurance. Likewise, they recommend that this time could be used with the presence of virtual “clinical case” classes, to maintain a constant practice of how to solve problems in a hospital environment and not forget prior knowledge. Finally, they suggest not continuing the tuition and monthly charges on the inmates until they return to the hospitals, because they do not benefit from the educational proposals of the universities. All the original answers, in Spanish, are found in the document “**Supplementary material**”.

**Figure 4.**
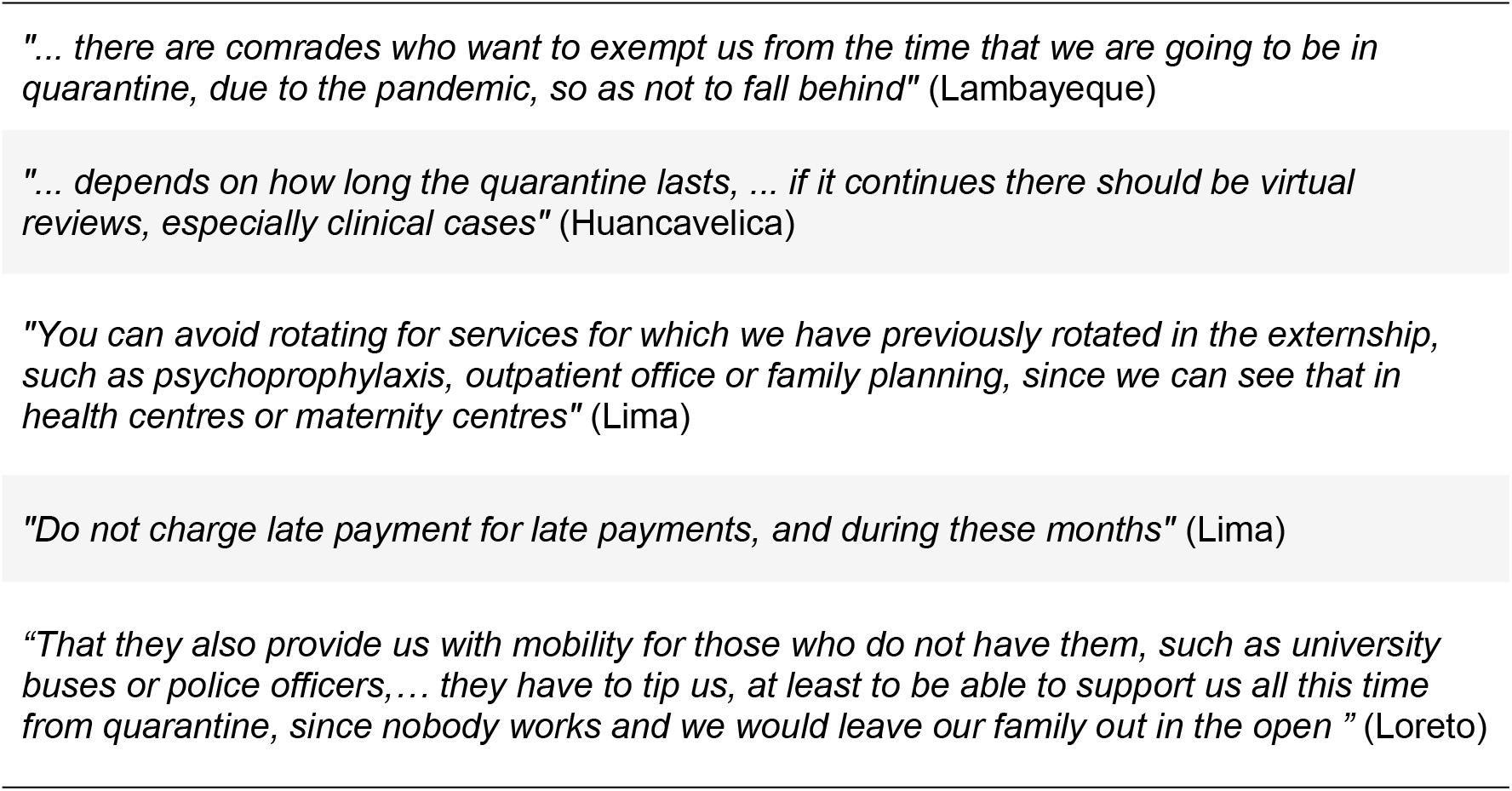
Solutions proposed by midwifery interns regarding the return of hospital practices in times of COVID-19

## DISCUSSION

Health personnel are a population exposed to the contagion of COVID-19. Although it is true that the inmates have not yet finished their university studies, the tasks they carry out and the risk of contagion can become the same as any hired worker (15,16). Even so, there are no guarantees in the delivery of personal protective equipment (PPE) and comprehensive insurance. Recent studies and the World Health Organization (WHO) agree that there is a global shortage of PPE, as well as medical furniture, mechanical ventilators, and hospital beds, which shows how this problem is replicated internationally (17-19)

On the other hand, the suspension of clinical practices and academic activities has substantially affected those students who were doing their internship in cities far from their origin, which has forced them to assume expenses that allow them to maintain their accommodation and food before the possibility of resume practice at any time. International evidence has reported this problem in those students who study abroad, where it is shown that they not only face the financial problem, but also a constant concern for their families in their cities of origin, being susceptible to anxiety and stress (20-22).

Regarding the perception of university management in a context of COVID-19, the midwifery interns report that in certain cases their coordinators have not commented on any definite position on the start of academic activities. This situation has been taking place in various countries. Studies carried out in China, where they have experienced a similar stage with SARS, suggest the implementation of virtual training on the current disease, maintain constant communication between students, teachers and administrative staff and, finally, manage remote psychological services for their student staff (2. 3); Likewise, the University of Washington suggests an active stance of academic institutions, where alliances are generated with local public entities that allow student healthcare support and scientific development (24).

One of the solutions proposed by the midwifery inmates was the reduction of the duration of the internship, due to the exposure time they could have in the hospital. We assume that the measure could be feasible depending on the evaluation of the missing time and the achievement of the competencies proposed for the boarding school. A previous study has established as a strategy that, in order not to postpone the wait, they begin with virtual classes on epidemiology, and then include the inmates in the disease control and prevention programs in first-level establishments, where there is a lower risk of contagion. (9)

The demand for the suspension of the monthly payment or other expenses towards the universities while the compulsory social immobilization lasts has been based on the student’s dissatisfaction with paying an incomplete or poorly delivered service; however, this attitude can be enhanced by the economic limitations inherent in the context, which escape the academic-hospital environment and also includes the loss of massive jobs and the increase in costs of certain foods due to panic, greatly limiting the economic income due to family (26).

It is necessary to consider that the results found are limited to the subjectivity of the midwifery student, with which it could be replicated in other fields of the health sciences or the perspective of the authorities, both hospital and university, could be evaluated. However, the strength of this study lies in the representativeness that has been sought, considering the presence of students from all regions of Peru.

Finally, we conclude that midwifery inmates perceive that the lack of personal protective equipment, the economic repercussions and the delay of procedures are the main problems where they have been affected. Likewise, this study reports solutions that could be considered for the next policies to be formulated.

## Data Availability

The data has been collected by the researchers themselves, they are not found in another repository.

## REFERENCES

1. Organización Mundial de la Salud. Preguntas y respuestas sobre la enfermedad por coronavirus (COVID-19). [Internet] New York: Organización Mundial de la Salud. [Cited: 22 Abril 2020] Available from: https://www.who.int/es/emergencies/diseases/novel-coronavirus-2019/advice-for-public/q-a-coronaviruses

2. Accinelli RA, Zhang-Xu CM, Ju-Wang JD, Yachachin-Chávez JM, Cáceres-Pizarro JA, Tafur-Bances KB, et al. COVID-19: La pandemia por el nuevo virus SARS-CoV-2. Rev Peru Med Exp Salud Publica. 2020;37(2). DOI: https://doi.org/10.17843/rpmesp.2020.372.5411

3. Organización Mundial de la Salud. Declaración conjunta de la ICC y la OMS: Un llamamiento a la acción sin precedentes dirigido al sector privado para hacer frente a la COVID-19. [Internet] New York: Organización Mundial de la Salud. [Cited: 22 April 2020] Disponible en: https://www.who.int/es/news-room/detail/16-03-2020-icc-who-joint-statement-an-unprecedented-private-sector-call-to-action-to-tackle-covid-19

4. Senado de la República de México. Implicaciones económicas de la pandemia por COVID-19 y opciones de política. [Internet] Ciudad de México: Dirección General de Finanzas. [Cited: 22 Abril 2020] Available from: http://www.bibliodigitalibd.senado.gob.mx/bitstream/handle/123456789/4829/NEcoronavirusimplicaciones%20econ%c3%b3micas%20010422020.pdf?sequence=1&isAllowed=y

5. Usher K, Bhullar N, Jackson D. Life in the pandemic: Social isolation and mental health. Journal of Clinical Nursing. 06 April 2020. DOI: https://doi.org/10.1111/jocn.15290

6. CEPAL. Las oportunidades de la Digitación en América Latina frente al COVID-19. [Internet] New York: Comisión Económica Para América Latina y el Caribe. [Cited 26 April 2020]. Available from: https://www.cepal.org/es/publicaciones/45360-oportunidades-la-digitalizacion-america-latina-frente-al-covid-19

7. Association of American Medical Colleges. Guidance on medical students participation in direct patient contact activities. [Internet] Washington: AAMC. [Cited: 26 April 2020] Available from: https://www.aamc.org/system/files/2020-04/meded-April-14-Guidance-on-Medical-Students-Participation-in-Direct-Patient-Contact-Activities.pdf

8. Association of American Medical Colleges. COVID-19: Updated Guidance for Medical Students’ Roles in Direct Patient Care. [Internet] Washington: AAMC. [Cited: 29 April 2020]. Available from: https://www.aamc.org/news-insights/press-releases/covid-19-updated-guidance-medical-students-roles-direct-patient-care

9. Bauchner H, Sharfstein. A Bold Response to the COVID-19 Pandemic: Medical Students, National Service, and Public Health. JAMA; 8 April 2020. DOI: https://doi.org/10.1001/jama.2020.6166

10. Diario Oficial El Peruano. Decreto Supremo que declara Estado de Emergencia Nacional por las graves circunstancias que afectan la vida de la Nación a consecuencia del brote del COVID-19. [Internet] Lima: Presidencia de la República. [Cited: 26 April 2020] Available from: https://busquedas.elperuano.pe/normaslegales/decreto-supremo-que-declara-estado-de-emergencia-nacional-po-decreto-supremo-n-044-2020-pcm-1864948-2/

11. Superintendencia Nacional de Educación Superior Universitaria. Sunedu e Indecopi exhortan a las universidades privadas a reprogramar matrículas, pagos o mensualidades. [Internet] Lima: SUNEDU. [Cited: 26 April 2020]

12. Instituto Internacional para la Educación Superior en Latinoamérica y el Caribe. COVID-19 y educación superior: De los efectos inmediatos al día después- [Internet] Caracas: IESALC. [Cited: 26 April 2020] Available from: http://www.iesalc.unesco.org/wp-content/uploads/2020/04/CQVID-19-060420-ES-2.pdf

13. Liang Z, Ooi S, Wang W. Pandemics and Their Impact on Medical Training. Academic Medicine. 2020 April. DOI: https://doi.org/10.1097/ACM.0000000000003441

14. Cleland JA. The qualitative orientation in medical education research. Korean Journal of Medical Education. 2017; 29(2): 61-71. DOI: https://doi.org/10.3946/kjme.2017.53

15. Eaves JL, Payne N. Resilience, stress and burnout in student midwives. Nurse Educ Today. 2019; 79(1): 188-193. DOI: https://doi.org/10.1016/j.nedt.2019.05.012

16. McCarthy B, Trace A, O’Donovan M, Brady-Nevin C, Murphy M, O’Shea M, O’Regan P. Nursing and midwifery students’ stress and coping during their undergraduate education programmes: An integrative review. Nurse Educ Today. 2018; 68(1): 197-209. DOI: https://doi.org/10.1016/j.nedt.2017.11.029

17. Swift A. COVID-19 and student nurses: a view from England. Journal of Clinical Nursing. 2020 April. DOI: https://doi.org/10.1111/jocn.15298

18. Fong ZV et al. Practical Implications of Novel Coronavirus COVID-19 on Hospital Operations, Board Certification, and Medical Education in Surgery in the USA. Journal of Gastrointestinal Surgery. 2020 April. DOI: https://doi.org/10.1007/s11605-020-04596-5

19. World Health Organization. Rational use of personal protective equipment for coronavirus disease 2019 (COVID-19). [Internet] New York: WHO. [Cited: 2 May 2020] Disponible en: https://apps.who.int/iris/bitstream/handle/10665/331215/WHQ-2019-nCov-IPCPPE_use-2020.1-eng.pdf

20. Cao W, Fang Z, Hou G, Han M, Xu X, Dong J, Zheng J. The psychological impact of the COVID-19 epidemic on college students in China. Psychiatry Research. 2020 May. DOI: https://doi.org/10.1016/j.psychres.2020.112934

21. Zhai Y, Du X. Mental health care for international Chinese students affected by the COVID-19 outbreak. The Lancet Psychiatry. 2020; 7(4). DOI: https://doi.org/10.1016/S2215-0366(20)30089-4

22. Astudillo-Pedroza M. Beneficios laborales específicos para los internos de medicina: Propuesta de un proyecto de ley. [Tesis] Lima: Universidad de Lima. [Cited el 03 de May del 2020] Available from: http://repositorio.ulima.edu.pe/bitstream/handle/ulima/3401/AstudilloPedrozaMartha.pdf?sequence=3&isAllowed=y

23. Meng L, Hua F, Bian Z. Coronavirus Disease 2019 (COVID-19): Emerging and Future Challenges for Dental and Oral Medicine. Journal of Dental Research. 2020 March. DOI: https://doi.org/10.1177/0022034520914246

24. Kim CS et al. One Academic Health System’s Early (and Ongoing) Experience Responding to COVID-19: Recommendations From the Initial Epicenter of the Pandemic in the United States. Acad Med. 2020 April. DOI: https://doi.org/10.1097/ACM.0000000000003410

25. Universidad Nacional Mayor de San Marcos. Plan curricular de la Escuela Profesional de Obstetricia, 2018. [Internet] Lima: EPO. [Cited el 02 de Mayo del 2020] Available from: https://medicina.unmsm.edu.pe/images/FacultadMedicina/EscuelaObstetria/plan-curricular-2018-epo.pdf

26. Nicola M et al. The Socio-Economic Implications of the Coronavirus and COVID-19 Pandemic: A Review. Int J Surg. 2020 April. DOI: https://doi.org/10.1016/j.ijsu.2020.04.018

